# Predicting skin cancer risk from facial images with an explainable artificial intelligence (XAI) based approach: a proof-of-concept study

**DOI:** 10.1101/2023.10.04.23296549

**Authors:** X. Liu, T.E. Sangers, T. Nijsten, M. Kayser, LM. Pardo, E.B. Wolvius, G.V. Roshchupkin, M. Wakkee

## Abstract

**Background:** Efficient identification of individuals at high risk of skin cancer is crucial for implementing personalized screening strategies and subsequent care. While Artificial Intelligence holds promising potential for predictive analysis using image data, its application for skin cancer risk prediction utilizing facial images remains unexplored. We present a neural network-based explainable artificial intelligence (XAI) approach for skin cancer risk prediction based on 2D facial images and compare its efficacy to 18 established skin cancer risk factors using data from the Rotterdam Study.

**Methods:** The study employed data from the Rotterdam population-based study in which both skin cancer risk factors and 2D facial images and the occurrence of skin cancer were collected from 2010 to 2018. We conducted a deep-learning survival analysis based on 2D facial images using our developed XAI approach. We subsequently compared these results with survival analysis based on skin cancer risk factors using cox proportional hazard regression.

**Findings:** Among the 2,810 participants (mean Age=68.5±9.3 years, average Follow-up=5.0 years), 228 participants were diagnosed with skin cancer after photo acquisition. Our XAI approach achieved superior predictive accuracy based on 2D facial images (c-index=0.72, SD=0.05), outperforming that of the known risk factors (c-index=0.59, SD=0.03).

**Interpretation:** This proof-of-concept study underscores the high potential of harnessing facial images and a tailored XAI approach as an easily accessible alternative over known risk factors for identifying individuals at high risk of skin cancer.

**Funding:** The Rotterdam Study is funded through unrestricted research grants from Erasmus Medical Center and Erasmus University, Rotterdam, Netherlands Organization for the Health Research and Development (ZonMw), the Research Institute for Diseases in the Elderly (RIDE), the Ministry of Education, Culture and Science, the Ministry for Health, Welfare and Sports, the European Commission (DG XII), and the Municipality of Rotterdam. G.V. Roshchupkin is supported by the ZonMw Veni grant (Veni, 549 1936320).

**Research in context:** *Evidence before this study:* We searched PubMed for articles published in English between Jan 1, 2000, and Sept 28, 2023, using the search terms “skin cancer” AND “artificial intelligence” OR “deep learning”. Our search returned more than 1,323 articles. We found no study had explored the feasibility of predicting the risk of developing skin cancer based on facial images that were taken before the first diagnosis of skin cancer. Although there were studies focused on deep learning image analysis and skin cancer, those are based on skin cancer lesion images. We found current skin cancer risk prediction models are still hampered by dependencies on complex patient data, including genetic information, or rely on self-reported patient data.

*Added value of this study:* In this study, we presented a neural network-based explainable artificial intelligence (XAI) approach for skin cancer risk prediction based on 2D facial images. To the best of our knowledge, our study is the first to utilize facial images as predictors in a skin cancer survival analysis. Our novel image-based approach showed superior performance when juxtaposed with traditional methods that relied on clinical and genetic skin cancer risk factors, as observed within our study population

*Implications of all the available evidence:* This proof-of-concept study underscores the high potential of harnessing facial images and a tailored XAI approach as an easily accessible alternative over known risk factors for identifying individuals at high risk of skin cancer.

## Introduction

Skin cancer, the most common form of cancer in individuals of European ancestry with lighter skin tones, presents a significant public health concern. The two most common types of skin cancer, basal cell carcinoma (BCC) and squamous cell carcinoma (SCC), together referred to as keratinocyte carcinoma (KC), account for an estimated 6 million^1^ new cases each year across the globe. BCC, the most prevalent variant, grows slowly, often appearing as nodular, pigmented, or waxy lesions on sun-exposed skin. SCC, though less common than BCC, is more aggressive and usually presents as a scaly or erythematous patch or nodule. Even less common, but far more lethal, is malignant melanoma, which causes an estimated 57,000 deaths globally each year.^2^ Due to longer life expectancies and past excessive UV exposure, the number of KC and melanoma cases has surged over recent decades, and this trend is anticipated to continue.^2^

Early diagnosis of skin cancer can mitigate morbidity and mortality by preventing the progression to late-stage disease.^3^ However, population-wide screening programs have not been proven cost-effective,^4^ or convincingly demonstrated a reduction in skin cancer-related mortality or morbidity. However, personalized screening programs for individuals at high risk of developing skin cancer^5^ are considered as a potentially more feasible strategy to combat the ongoing skin cancer epidemic.^6^ Although accurate stratification of individuals at increased risk of developing skin cancer is essential for targeted screening, it remains a challenge to develop tools with sufficient predictive performance that are suitable for wide-scale use. Many existing prediction^7,8,9^ models are hampered by dependencies on complex patient data, including genetic information, or rely on self-reported patient data, thereby making them susceptible to recall bias and social-desirability bias.^10^

Artificial intelligence (AI), specifically convolutional neural networks (CNNs), have demonstrated high accuracy in detecting skin cancer from skin lesion images in recent years.^11^ Yet, their potential in stratifying individuals based on skin cancer risk remains largely uncharted due to the scarcity of longitudinal datasets. One potential direction for such screening strategies involves the use of personal facial images. These images could reveal key risk factors associated with skin cancer, such as age, skin color, and signs of UV damage. Furthermore, capturing facial images requires minimal effort (such as taking a selfie with a smartphone) and is not affected by recall bias thus offering a potentially easily accessible tool for identifying high risk individuals in the general population.

Here, we develop a neural network-based explainable artificial intelligence (XAI)^36,37^ approach that predicts skin cancer risk from 2D-facial images and utilize population-based data from the Rotterdam Study (RS) to demonstrate its performance. To assess its effectiveness, we compared the performance of our novel image-based approach with traditional clinical and genetic skin cancer risk factors in RS.

## Methods

### Study design and participants

The RS is an ongoing large prospective population-based cohort study in Ommoord, a region in Rotterdam, the Netherlands.^12^ Since January 1990, the RS has been enrolling individuals aged 50 and over from the general population. Participants undergo comprehensive baseline examinations and visit a dedicated study center every 3-4 years. During these visits they also undergo a full-body skin examination (FBSE) performed by a dermatology-trained physician which also focuses on detecting skin (pre-)malignancies and skin cancer. As of 2010, standardized full facial images are taken of participants using a Premier 3dMD face3-plus UHD camera (3dMD Inc., Atlanta, GA, USA).

### Skin cancer and risk factor measurements

Skin cancer diagnoses, both prior to and after the facial photo was taken, along with their respective body locations, were collected for all RS participants by linkage to the Dutch nationwide network and registry of histo– and cytopathology (PALGA).^13^ Skin cancer-related risk factors were collected through a combination of home interviews and study center visits. Available skin cancer determinants included sex, age, skin color, hair color, eye color, pigment status (combined variable^14,15^ hair and eye color), number of naevi, baldness in men, body mass index (BMI), socioeconomic status, history of living in a sunny country, tendency to develop sunburn, alcohol intake, coffee consumption, smoking, Glogau wrinkle classification, and a genetic risk score (GRS) for KC as well as melanoma utilizing single-nucleotide polymorphisms (SNPs) that are significantly associated with these specific types of skin cancer.^34,35^

### Case definition

***Events*** were defined as participants, who received their first diagnosis of skin cancer (BCC, SCC or melanoma) after the date of the facial images being taken. To ensure the exclusion of participants who already had a skin cancer diagnosis at the time of the photograph (i.e., ***left-censored*** samples), individuals with a confirmed histopathological skin cancer diagnosis prior to their photograph or within 30 days after the FBSE were excluded from the study. Follow-up of all participants ended at the time of death, or the date of censoring on July 1^st^, 2018, whichever came first. Death was ascertained through linking with the municipal register. ***Right-censored*** samples were defined as participants, who had not received a diagnosis of skin cancer by the date of censoring (July 1st, 2018), or who died before the date of censoring.

### Facial image acquisition and preprocessing

Facial images of the RS participants were taken using a 3dMDface system (3dMD Inc., Atlanta, GA, USA) photogrammetric device by medical doctors, who were specifically trained in operating the device. The system comprised a central modular camera unit, flanked by two additional side units, and underwent daily calibration. Image acquisition took place in a designated 3D imaging room with consistent ambient lighting. An adjustable chair was used in a fixed position, to ensure a standard level of height and fixed distance between subjects and the camera system. Participants were requested to remove glasses and to wear a hair band to prevent hair from obscuring the forehead or ears. During the image capture process, participants faced the central modular camera unit maintaining a neutral facial expression with their eyes open. Frontal 2D facial images were automatically derived from the 3dMD software.

For the detection of facial landmarks, we utilized Dlib,^16^ a facial image processing library. The detected facial landmarks were subsequently employed to crop and align the facial regions, as illustrated in Figure S8. To exclude the neck or shoulder regions from the images, those areas were masked as black pixels. Additionally, histogram equalization was applied to enhance the visual quality of each facial image as well as to mitigate potential lighting variations. The final resolution employed for our analysis was set to 224×224 pixels.

### Deep learning analysis

#### AI-based endophenotypes derived from facial images

An autoencoder is an artificial neural network architecture employed for learning compressed representations of input data in a self-supervised manner, meaning that it does not require patient-specific information during the training process. The autoencoder consists of an encoder and a decoder, which collectively enable non-linear feature mapping. The encoder is responsible for compressing high-dimensional facial images into low-dimensional latent features, while the decoder reconstructs facial images from these latent features.

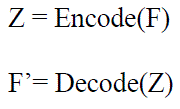

where Z = [Z_0_, Z_1_, …, Z_N_] denotes the N latent features, Encode() and Decode() functions correspond to the down-sampling and up-sampling processes F denotes the input 2D facial image, while F′ represents the reconstructed 2D facial image.

For our analysis, we used a deep convolutional autoencoder^17^ (detailed architecture in Figure S3c) consisting of four encoder layers and four decoder layers to derive low-dimensional representations, which we further defined as facial endophenotypes. To make the trade-off between reconstruction error and dimensional complexity, we conducted experiments with varying numbers of endophenotypes. The optimum number was found to be 200. The derived facial endophenotypes were further used as predictors in the survival analysis.

#### Explainable AI (XAI) techniques

By selectively decoding a subset of endophenotype(s), we are able to generate a sequence of facial images showing changes in facial features corresponding to the changes of selected endophenotype(s). Implementation details about the selectively decoding can be found in Figure S5a (in supplementary).

#### Cox proportional hazard regression (CPH) analysis

We performed a survival analysis employing cox proportional hazard regression (CPH),^18^ to predict the risk of participants developing skin cancer over time. Predictors included either the 18 risk factors or the 200 facial endophenotypes. Additionally, time-to-event (TTE) was included as an essential predictor in the model training. For events, TTE was calculated as the interval between the date of the first diagnosis of skin cancer and the date when the facial image was taken; In the case of right-censored samples, TTE was calculated as the interval between the date of censoring (or death) and the date when a facial image was taken.

#### Deep cox proportional hazards regression (DCPH) analysis

A deep cox proportional hazard regression (DCPH) model is an extension of the linear cox proportional hazards (CPH) model. Compared to DPH, DCPH enables non-linear modeling of the predictors, and thus it is able to model more complex relationships between predictors and the risk.^19^ The parameters of the DCPH model are optimized by minimizing the following log-partial likelihood cost function which is widely used in survival analysis models:^20,21,22^

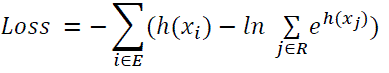

where E represents the set of events and R denotes the set of right-censored samples, and the TTE for individual j is greater than the TTE for individual i, x_i_ and x_j_ represent the predictors for individuals i and j, and h(x) and h(x_j_) represent the network predictions for individuals i and j. The DCPH model consisted of two fully connected layers and one neuron in the output layer. Detailed architectures of the DCPH model can be found in Supplementary Figure S3b.

#### Deep Convolutional cox proportional hazards regression analysis (DCCPH)

Integrating 2D convolutional neural networks^23^ with the cost function of DCPH, we can utilize the entire facial image as input and directly compute a skin cancer risk score for each participant. This integrated model was referred to as the deep convolutional cox proportional hazard (DCCPH) model, which consisted of four convolutional layers, two fully connected layers and one neuron in the output layer. Detailed architectures of the DCCPH model can be found in Supplementary Figure S3c.

### Survival analysis and experiment settings

In all experiments of the survival analysis, left-censored samples (i.e., participants diagnosed with skin cancer prior to the facial image being taken) were excluded. We utilized three models in the survival analysis: CPH, DCPH, and DCCPH. For both the CPH and DCPH models, we explored three types of predictors: 1) age alone, 2) 18 known risk factors, and 3) facial endophenotypes. In the DCCPH model, we employed 2D facial images without extracting facial endophenotypes as predictors.

All the three models estimate the risk over time, but we only provide the risk score at a single time point, which is at the end of the study. To evaluate the predictive performance of the models, we calculated the concordance-index (c-index) using the following equation:

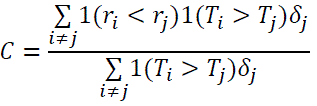

where δ_j_ is a variable that is assigned a value of 1, if individual j is an event and 0 if individual j is a right-censored sample, T is the TTE for an individual and r is the risk score for an individual. A c-index of 0.5 indicates a random prediction, while a c-index of 1 indicates a perfect prediction. In all prediction experiments, we employed a consistent practice of splitting the samples into a training set and a test set, with an 80:20% ratio. We repeated this split 20 times with random permutations and calculated the mean and standard deviation of the c-index.

We performed an additional analysis where we stratified the right-censored samples into different subgroups based on their follow-up years. It is important to note that patient information in this study was not updated beyond July 1st, 2018. Consequently, some right-censored samples had a relatively short follow-up period and may have been diagnosed with skin cancer shortly after that date. These right-censored samples with a shorter follow-up time could be deemed less reliable compared to those with longer follow-up periods. In consequence, including right-censored samples with short follow-up years in the analysis could potentially impact the prediction model negatively. Therefore, we stratified the right-censored samples into 5 subgroups based on the follow-up years, while ensuring age matching among the subgroups. The subgroups were defined as follows: 1) > 2 follow-up years (N=1139); 2) > 3 follow-up years (N=1129); 3) > 4 follow-up years (N=840); 4) > 5 follow-up years (N=548); 5) > 6 follow-up years (N=535), The event group consisted of participants who were diagnosed with skin cancer at any location on the body (N=228). Subsequently, we performed separate survival analysis for each sub-group in this additional analysis.

### Ethical considerations

The Rotterdam Study has been approved by the Erasmus MC Medical Ethical Committee (MEC-02-1015), and by the Dutch Ministry of Health, Welfare and Sport (Population Screening Act, reference 3295110-1021635-PG). Written informed consent was obtained from all participants.

## Results

### Study population and risk factors

We included a total of 3,371 participants from the RS cohort, who had a 2D-facial image and underwent assessment for skin cancer risk factors. Among them, 23.4% (n=789) were diagnosed with skin cancer, of which 71.1% (n=561, ***left-censored*** participants) were diagnosed prior to the facial image being taken, and 28.9% were diagnosed afterwards (n=228, ***events***). Figure S7 shows the histogram distribution of time-to-event (TTE) of these 228 events, with an average TTE of 971 days (SD 678 days). Among these 228 events, 163 had a BCC, 68 had an SCC and 11 had a melanoma. Left-censored participants were excluded from the survival analysis. Table 1 shows the distribution of the 18 potential skin cancer risk factors among the study population (N=2,810), of which the median age was 67.1 years (IQR 14.1 years) and 57.4% were women. We examined the association between known skin cancer risk factors and the facial endophenotypes we derived from the facial images (Figure S4), and visually represented the identified associations on the face (Figure S5) via XAI techniques.

**Table 1:** Baseline characteristics of the RS study population.

|  | Participants, who did not develop skin cancer (Right-censored samples)<br>N=2582 | Participants, who developed skin cancer after facial image taken (Events)<br>N=228 |
| --- | --- | --- |
| Age at FBSE in years, median (IQR) | 66.8 (14.2) | 70.6 (12.0) |
| Sex |  |  |
| -Women (%) | 1484 (57.5%) | 129 (56.6%) |
| -Men (%) | 1098 (42.5%) | 99 (43.4%) |
| BMI<br>mean (SD) | 27.67 (4.39) | 27.75 (3.97) |
| **Skin color**<br>- Olive color-light brown (%)<br>- White (%)<br>- Pale (%) | 373 (14.4%)<br>1908 (73.9%)<br>301 (11.7%) | 23 (10.1%)<br>183 (80.3%)<br>22 (9.6%) |
| **Hair color when young**<br>-Black (%)<br>-Brown/dark blonde (%)<br>-Light blonde (%)<br>-Red (%) | 155 (6.0%)<br>1844 (71.4%)<br>505 (19.6%)<br>78 (3.0%) | 9 (3.9%)<br>166 (72.8%)<br>40 (17.5%)<br>13 (5.7%) |
| **Eye color**<br>-Brown (%)<br>-Intermediate (%)<br>-Blue (%) | 596 (23.1%)<br>211 (8.2%)<br>1775 (68.7%) | 48 (21.1%)<br>21 (9.2%)<br>159 (69.7%) |
| **Pigment status (combined variable<sup>14,15</sup>**<br>**hair and eye color)**<br>-Light (%)<br>-Intermediate (%)<br>-Dark (%) | 538 (20.8%)<br>1467 (56.8%)<br>577 (22.3%) | 49 (21.5%)<br>134 (58.8%)<br>45 (19.7%) |
| **Baldness**<br>-No/almost no (%)<br>-Mild (%)<br>-Severe (%) | 1923 (74.5%)<br>346 (13.4%)<br>313 (12.1%) | 157 (68.9%)<br>31 (13.6%)<br>40 (17.5%) |
| **Number of naevi**<br>- >=100 (%)<br>- 50-99 (%)<br>- 25~49 (%)<br>- <25 (%) | 49 (1.9%)<br>141 (5.5%)<br>371 (14.4%)<br>2021 (78.3%) | 4 (1.8%)<br>9 (3.9%)<br>30 (13.2%)<br>185 (81.1%) |
| **Glogau wrinkle classification**<br>- 1 and 2 (%)<br>- 3 (%)<br>- 4 (%) | 207 (8.0%)<br>2027 (78.5%)<br>348 (13.5%) | 12 (5.3%)<br>175 (76.8%)<br>41 (18.0%) |
| **Socioeconomic status**<br>-high (%)<br>-medium (%)<br>-low (%) | 755 (29.2%)<br>1505 (58.3%)<br>322 (12.5%) | 56 (24.6%)<br>134 (58.8%)<br>38 (16.7%) |
| **History of living in a sunny country**<br>-Yes (%)<br>-No (%) | 150 (5.8%)<br>2432 (94.8%) | 17 (7.5%)<br>211 (93.5%) |
| Tendency to develop sunburn<br>-Yes (%)<br>-No (%) | 866 (33.5%)<br>1716 (66.5%) | 86 (37.7%)<br>142 (62.3%) |
| Alcohol intake<br>g/day<br>mean (SD) | 7.95 (8.57) | 8.08 (9.42) |
| Smoking<br>-Ever (%)<br>-Never (%) | 1756 (68.0%)<br>826 (32.0%) | 157 (68.9%)<br>71 (31.1%) |
| Coffee consumption (cups/day)<br>mean (SD) | 3.03 (1.83) | 2.99 (1.81) |
| GRS_KC<br>median (SD) | 1.03 (0.25) | 1.06 (0.27) |
| GRS_MM,<br>median (SD) | 7.03 (0.47) | 7.06 (0.46) |

### Skin cancer risk prediction in survival analysis

Table 2 shows the c-index values for different prediction models using various predictors as input. In the risk factor analysis, the age-only analysis yielded a c-index of about 0.55, which increased to 0.59 when all other known risk factors were included. However, the analysis of facial endophenotypes resulted in higher c-index values of 0.72, which remained similar at 0.71 when using the facial images without extracting the facial endophenotypes. Comparing the prediction models, the CPH showed better performance when using risk factors as input, while the DCPH was more effective when employing facial endophenotypes as input.

**Table 2:** C-index (mean/SD) of skin cancer risk prediction in survival analysis.

| Method (predictors) | Skin cancer on any location of body<br>N = 228 | Skin cancer on face<br>N = 136 | Skin cancer on other parts of the body except face<br>N=113 |
| --- | --- | --- | --- |
| CPH age only | 0.550/0.048 | 0.554/0.036 | 0.543/0.051 |
| CPH 18 known risk factors | 0.589/0.034 | 0.586/0.047 | 0.595/0.060 |
| DCPH 18 known risk factors | 0.572/0.044 | 0.520/0.067 | 0.532/0.069 |
| CPH facial endophenotypes | 0.685/0.033 | 0.693/0.037 | 0.674/0.054 |
| DCPH facial endophenotypes | 0.721/0.045 | 0.732/0.058 | 0.718/0.049 |
| DCPH facial endophenotypes + age | 0.723/0.039 | 0.738/0.060 | 0.721/0.062 |
| DCCPH 2D facial image without extracted endophenotypes | 0.713/0.041 | 0.721/0.051 | 0.703/0.049 |

CPH: Cox proportional hazard regression
DCPH: Deep Cox proportional hazard regression
DCCPH: Deep convolutional cox proportional hazards regression
Only Right-censored samples (N= 535) with > 6 follow-years were included.
Results were based on imputed risk factors. Table S2 shows the results based on non-imputed risk factors.

In our main analysis, we focused on skin cancer occurring at any location on the body. We further stratified the event group into two subgroups: skin cancer on the face and skin cancer on other parts of the body excluding the face. Interestingly, we observed similar patterns and trends in the analysis across different stratifications based on the location of skin cancer.

### Facial endophenotypes association with known risk factors

Figure 1 shows the associations between traditional skin cancer risk factors and statistically significant facial endophenotypes identified in the survival analysis (CPH facial endophenotypes) in Table 2. Combining the effects of 12 statistically significant facial endophenotypes, several key patterns emerged. Strong associations of facial endophenotypes were observed with age (positive), Glogau wrinkle classification (positive), history of living in a sunny country (positive), BMI (negative), and hair color and pigment status (higher risk in lighter color). Weaker associations were found with sex (higher risk in males), tendency to develop sunburn (negative) and skin color (higher risk with lighter skin color). No significant associations were detected with alcohol consumption, coffee consumption, smoking, eye color, social economic status, number of naevi, baldness and genetic risk score.

**Figure 1.**
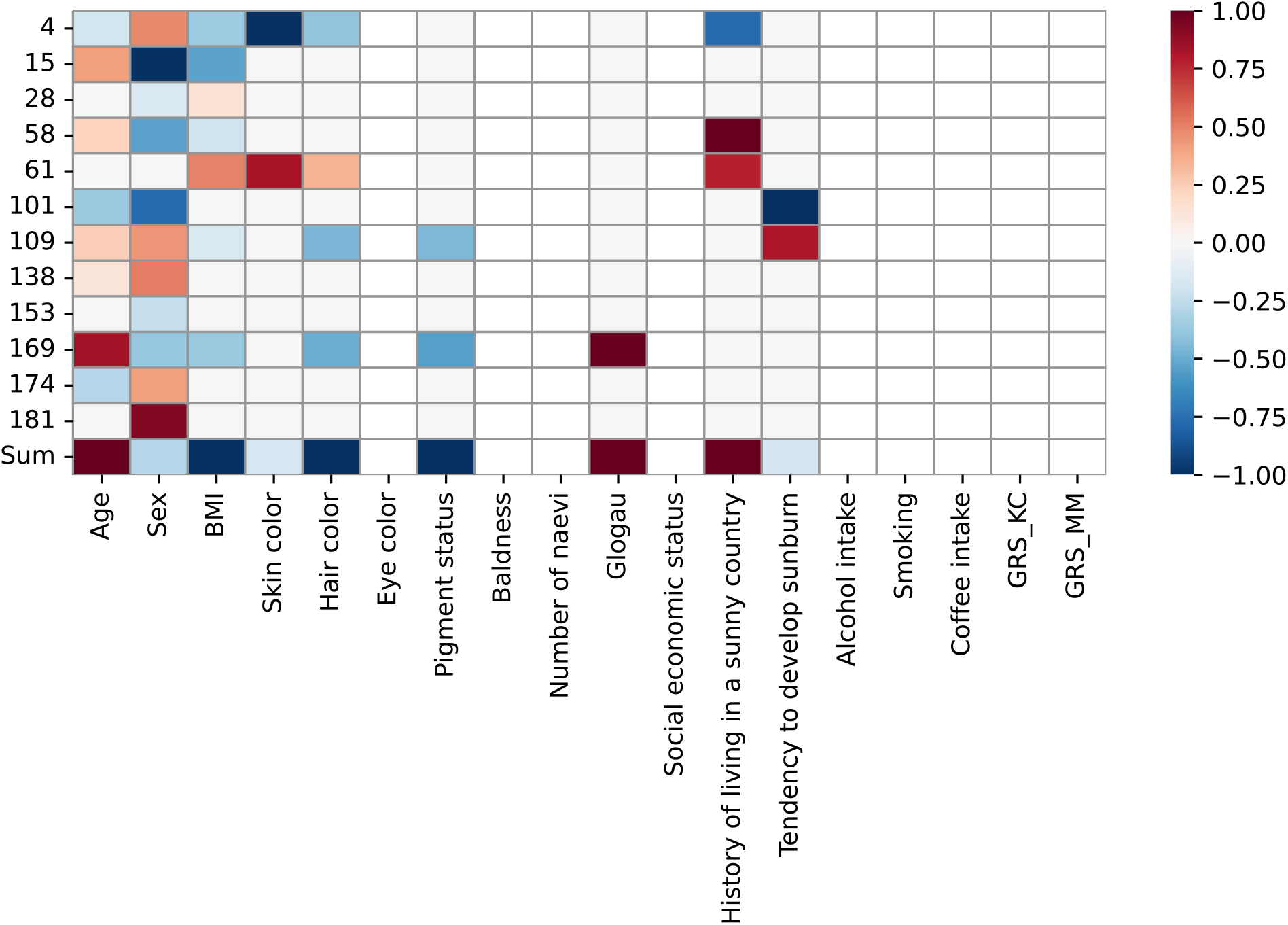
Statistically significant facial endophenotypes in survival analysis are associated with known risk factors, in the analysis of skin cancer on any location of the body. The x-axis represents the different risk factors, while the y-axis represents the index of facial endophenotypes that exhibited statistical significance in the survival analysis. The “sum” row represents the summation of each row. The values of associations are normalized from –1 to 1 for each column, where red indicates a positive association, indicating a higher risk of skin cancer associated with higher values of the corresponding risk factor, such as age.

### Explainable AI (XAI)

In order to gain insights into the facial features contributing to skin cancer prediction, we selectively decoded the statistically significant facial endophenotypes identified in the survival analysis (CPH facial endophenotypes) and generated a sequence of facial images representing low to high risk of skin cancer via XAI techniques (implementation details in Figure S5a). Figure 2 presents synthesized facial images that the prediction model considered as having low to high risk of skin cancer, stratified based on whether the cancer occurred on the face, on other body parts excluding the face, or anywhere on the body including the face. The result indicates that factors such as a lower BMI or increased facial erythema might be linked to a higher risk. Notably, participants in the sub-group of skin cancer occurring outside the face had never been diagnosed with skin cancer on the face before the date of censoring.

**Figure 2:**
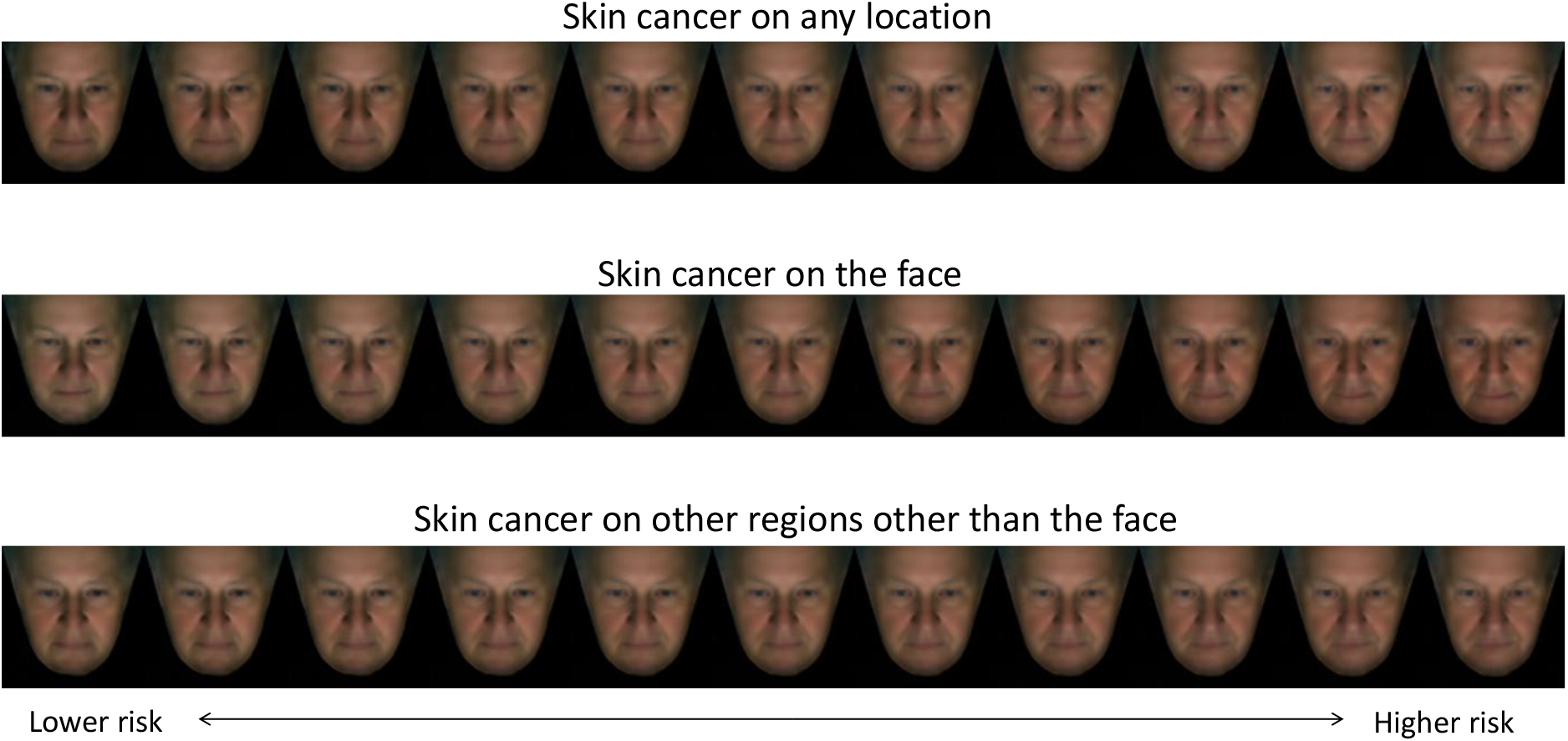
Reconstruction of faces representing lower to higher risk for developing skin cancer on the face, and other body locations excluding the face, as well as anywhere on the body including the face. The synthetic faces shown in the figure were reconstructed via selectively decoding statistically significant facial endophenotypes in the survival analysis (CPH facial endophenotypes) in Table 2.

### In-depth analysis of TTE in survival analysis

The relationship between the skin cancer prediction score and TTE of events during the test stage of the survival analysis (CPH and DCPH facial endophenotypes) is illustrated in Figure 3. Notably, the TTE information of each participant was not included into the model as a predictor in the test stage. The plot reveals that events with a higher prediction score, indicating a higher risk of skin cancer, tend to have a shorter TTE. A shorter TTE implies that participants were diagnosed with skin cancer earlier after the facial images were taken, suggesting that their faces may have exhibited early signs of skin cancer at the time the photo was made. The CPH and DCPH models effectively identified these participants, assigning them to overall higher risk scores compared to subjects with a longer TTE. This effect, although to a lesser extent, was also observed for participants with skin cancer on other parts of the body excluding the face (Figure S9).

**Figure 3:**
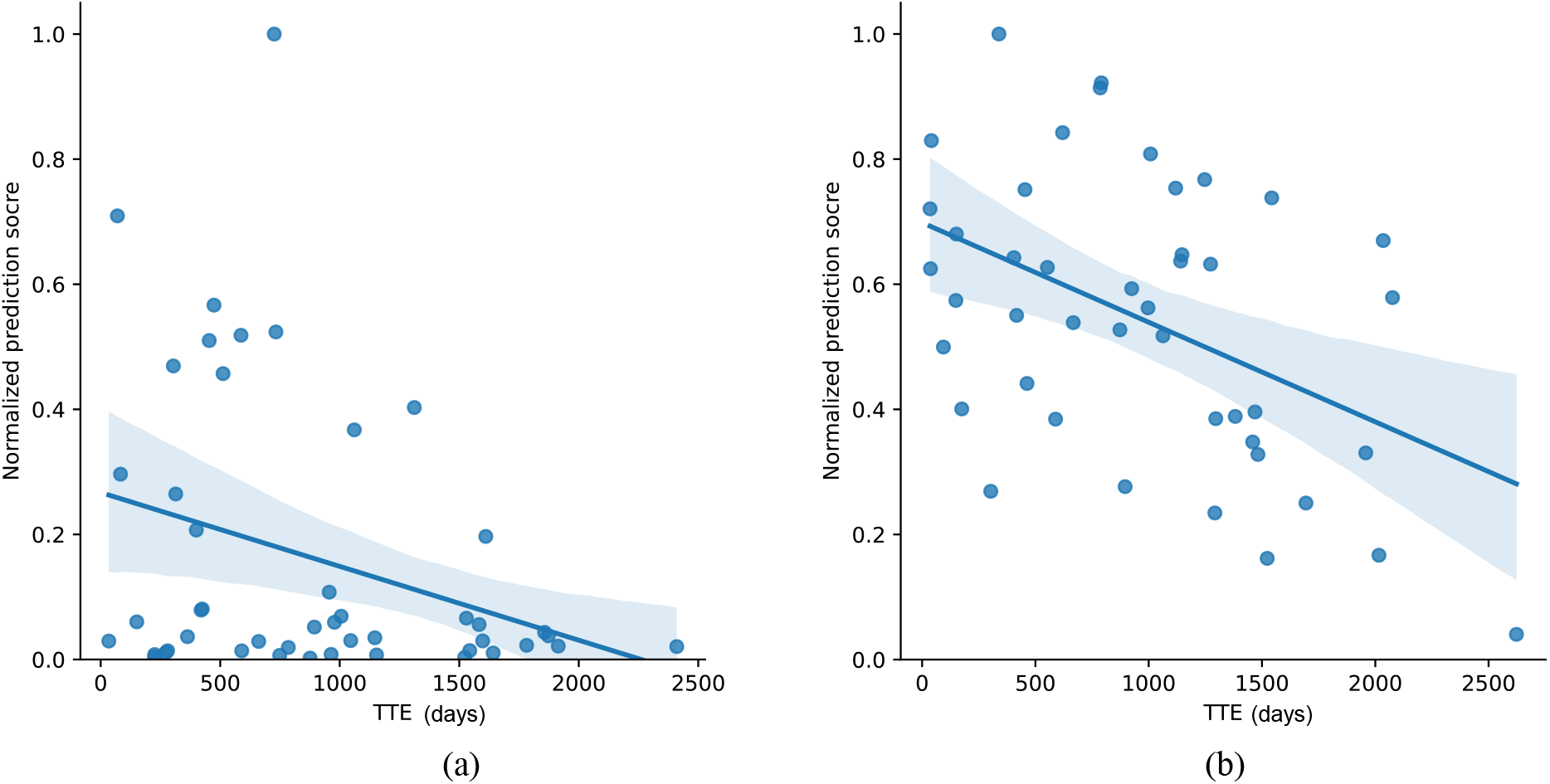
Relationship between risk prediction score and time to event (TTE) for N=44 events in the test set. a) Analysis of (CPH facial endophenotypes) and b) Analysis of (DCPH facial endophenotypes) for skin cancer at any location on the body. The X-axis represents the TTE in days, while the Y-axis represents the prediction risk score normalized to a range of 0 to 1. A higher predicted score indicates a higher risk of skin cancer.

Figure 4 shows the survival analysis results with the stratification of right-censored samples based on their follow-up years. Age was matched between the event group and right-censored subgroups, so the c-index curve of (CPH age) served as a baseline. It is evident that compared to (CPH age), the results of (CPH 18 known risk factors), (CPH facial endophenotypes) and (DCPH facial endophenotypes) showed a trend of increasing c-index in subgroups with longer follow-up years.

**Figure 4:**
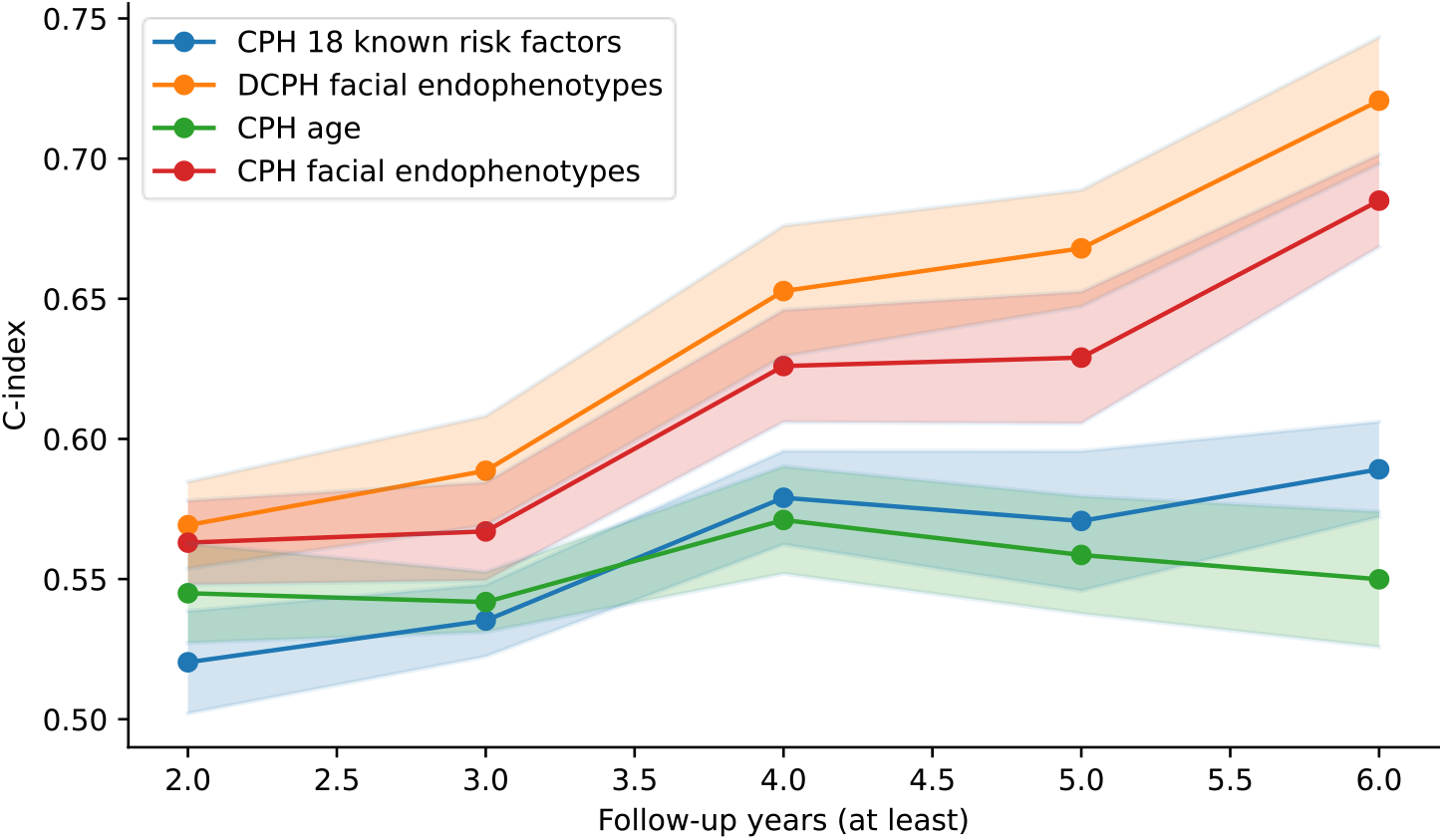
Stratification of right-censored samples based on follow-up years. The solid line represents the mean result, while the transparent surrounding area denotes the standard deviation. (CPH 18 known risk factors): Results obtained from a CPH model using risk factors as predictors. (DCPH facial endophenotypes): Results obtained from a DCPH model using facial endophenotypes as predictors.

## Discussion

Our research underscores the promise of using facial endophenotypes, extracted from 2D facial images through AI techniques, as indicators for developing skin cancer in the future. We demonstrate that the innovative use of facial image-based predictions outperformed traditional methods relying on many known risk factors, which are normally identified based on complex patient information, including extensive questionnaires, genetic data, and clinical parameters identified during physical skin cancer screening, such as nevi count, AK, or wrinkles. Our analysis confirms the robust relationship between these facial endophenotypes and established skin cancer risk determinants. We found a notable correlation between the most predictive facial endophenotypes and certain recognized risk factors for skin cancer.

Facial endophenotypes provide a rapid, user-friendly and explainable AI-based alternative to existing risk factors, particularly when comprehensive patient information is difficult to collect on a large scale. Furthermore, these endophenotypes might offer a more consistent prediction of skin cancer risk than relying on questionnaire data, sidestepping potential recall biases. To the best of our knowledge, our study is the first to utilize facial images and facial endophenotypes derived from these images as predictors in a skin cancer survival analysis.

### Relevance to existing literature

Previous modeling studies reported targeted skin cancer screening of high-risk populations to be a cost-effective^24^ population-based intervention to reduce skin cancer related morbidity and mortality. However, the characteristics of individuals that are considered ‘high risk’ vary across studies.^25^ Our findings provide promises that facial endophenotypes can help in narrowing the scope of high-risk individuals to optimize cost-effectiveness and improve the feasibility of targeted screening programs.

The current literature on skin cancer risk prediction encompasses a range of prediction models with varying levels of accuracy. While some models have demonstrated promising results using relatively simpler factors, such as the number of actinic keratosis and coffee consumption (c-index 0.6),^26^ others have incorporated more extensive patient information to achieve improved accuracy. Notably, a model focused on melanoma risk achieved c-index values of 0.72 for any melanoma and 0.69 for invasive melanoma by incorporating seven relevant predictors.^27^ In our study, utilizing facial endophenotypes outperformed the predictive accuracy based on known risk factors alone, yielding a c-index of 0.73, which surpasses the performance of many published models. Recent research has broadened its scope, incorporating up to 32 genetic and non-genetic risk factors to remarkably enhance the accuracy of predicting future skin cancer development.^40^ Nonetheless, it’s vital to acknowledge that making direct comparisons between our study and prior investigations presents challenges due to variations in the populations, research methodologies employed, and, notably, the divergent primary outcomes assessed in these studies.

Our study also revealed associations between several facial endophenotypes and known risk factors for skin cancer such as age, sex, BMI, Glogau wrinkle score, hair and skin color, history of living in a sunny country, tendency to develop sunburn, and pigment status. Age is a well-known risk factor for skin cancer,^28,29,30^ with many existing prediction models relying heavily on it. Similarly, our prediction model leaned heavily on age as a discriminative variable when our analysis was restricted to known skin cancer risk factors.

Interestingly, we were also able to corroborate previous studies that found an inverse correlation between BMI and KC, meaning a higher BMI appears to reduce the risk of KC).^31,32^ However, other studies focusing on melanoma observed a positive correlation with BMI.^33^ Given the substantial proportion of KC patients within our study cohort relative to melanoma patients, this has likely skewed our results towards an inverse correlation.

Furthermore, we found several facial endophenotypes associated with sex to be predictive of skin cancer. Men are known to have a higher risk of developing skin cancer than women, particularly in the case of melanoma. This disparity can be attributed to a mix of genetic and behavioral factors, including men’s tendency to have more sun exposure and less protection from ultraviolet (UV) radiation due to occupational and recreational activities and less care to protect from UV with cosmetics. Moreover, individuals with lighter skin tones and hair colors are more susceptible to skin cancer, including both KC and melanoma, due to their decreased melanin levels, which leaves them less protected against UV radiation. Residing in a sunny country also increases skin cancer risk due to higher cumulative exposure to UV radiation. Surprisingly, our study identified an inverse relationship between the tendency to develop sunburn and the risk of skin cancer. This counterintuitive finding could be attributed to a chance finding as this association was relatively weak. Furthermore, we were unable to correlate other known skin cancer risk factors to facial endophenotypes, such as coffee consumption and the number of nevi. This outcome is anticipated, as these risk factors would be less likely to be reflected in facial images.

### Strengths and limitations

Strengths of this study include the use of extensive population-based data from the Rotterdam Study, with over 7 years of follow-up data. Additionally, the inclusion of nationwide data on skin cancer diagnosis from the Dutch National Histopathology Registry reduces the risk of having missed skin cancer diagnoses in the studied individuals during the study follow-up period. Nevertheless, the results of our study should be understood in the context of several limitations. First, given the proof-of-concept design of our study and, as far as we are aware of, no publicly available datasets worldwide containing both facial images and follow-up skin cancer data, we were unable to externally validate our algorithm using an independent dataset. Second, facial images in the Rotterdam Study were collected using a highly standardized method with a dedicated camera setup, which could result in suboptimal performance when other, out-of-distribution facial photos taken under different capturing conditions (e.g., selfies taken with a front-facing smartphone camera) are used in our AI model, which can be tested in future studies. Also, a portion of right-censored samples included in our analysis had a short follow-up period, meaning they might have developed skin cancer shortly after this study’s follow-up period ended, potentially impacting the prediction model negatively. Furthermore, as Rotterdam Study participants are of Dutch European ancestry and thus in majority having fair skin colour, the training and testing dataset of this study was unbalanced in terms of skin colour diversity, which may make our model less reliable when used for individuals with darker skin tones. Moreover, the study cohort is relatively old (mean age around 68) affecting the generalizability of the findings, suggesting that the impact of age may be even more dominant in the general population with a different age distribution, which should be tested in the future. Finally, we implemented an XAI approach in an attempt to shed light on the risk predictions made by our AI system. We did this by generating synthetic facial images associated with higher or lower risks of developing skin cancer. This method provides some insights, indicating that factors such as a lower BMI or increased facial erythema might be linked to a higher risk. But the inherent complexity of deep learning algorithms restricts our ability to fully understand the specific elements that boost the model’s accuracy.

## Conclusion

In conclusion, our research underscores the high potential of using facial images in deep-learning models for targeted skin cancer screening. The novel AI-driven approach we introduce here eliminates the need for collecting information via lengthy questionnaires, DNA collection, genotyping, or in-person evaluations. However, before it can be integrated into personalized screening programs, further validation within diverse population samples and less standardized setting is needed.

## Data Availability

De-identified data used in this study is currently not publicly available. Researchers interested in data access should contact the corresponding author. Data requests will need to undergo ethical and legal approval by the relevant institutions.

https://gitlab.com/xianjingliu/ai_skin_cancer_risk_prediction

## Acknowledgements

The Rotterdam Study is funded by the Erasmus Medical Center and Erasmus University, Rotterdam, Netherlands Organization for the Health Research and Development (ZonMw), the Research Institute for Diseases in the Elderly, the Ministry of Education, Culture and Science, the Ministry for Health, Welfare and Sports, the European Commission (Directorate-General XII), and the Municipality of Rotterdam. G.V. Roshchupkin is supported by the ZonMw Veni grant (Veni, 549 1936320). The authors are grateful to the study participants, the staff from the Rotterdam Study, and the participating general practitioners and pharmacists.

## Author contributions

All authors made significant contributions to this scientific work and approved the final version of the manuscript. X.L. and T.E. were involved in the conception and design of the study, conducted the method development and experiments, and wrote the article. M.W and G.V.R. were involved in the conception and design of the study, supervised the method development and design of experiments, and co-wrote the article. T.N, M.K, LM.P. and E.B.W. were involved in the conception and design of the study, reviewed the manuscript, and provided consultation regarding the analysis and interpretation of the data. X.L. and T.E. have directly accessed and verified the underlying data reported in the manuscript.

