## Supplementary for "Predicting skin cancer risk from facial images with an explainable artificial intelligence (XAI) based approach: a proof-of-concept study"


| Content | Page number |
|---|---|
| 1. Data characteristics of the non-imputed risk factors in the study population. | 2-4 |
| 2. Survival analysis prediction c-index based on imputed or non-imputed risk factors. | 5 |
| 3. Survival analysis based on all right-censored samples, or right-censored sample > 6 follow-up years. | 6-7 |
| 4. Detailed architectures of the deep learning models used in this study | 8-9 |
| 5. Associations analysis between facial endophenotypes and skin cancer risk factors | 10 |
| 6. Implementation details of our explainable artificial intelligence (XAI) techniques | 11-14 |
| 7. Binary prediction between right-censored participants and participants ever diagnosed with skin cancer | 15 |
| 8. Distribution of the Time-to-event of events | 16 |
| 9. Preprocessing of the 2D facial images | 17 |
| 10. Relationship between risk prediction score and time to event, for skin cancer at the body other than the face | 18 |

#### 1. Data characteristics of the non-imputed risk factors in the study population.

Table S1: Data characteristics of the study population. Risk factors were not imputed.

| | Right-censored samples | Events | | Light-censored samples | |
|---|---|---|---|---|---|
| | (N=2117) | (N=184) | p-value two-sample t-test with right-censored samples | (N=482) | p-value two-sample t-test with right-censored samples |
| Age at FBSE in years, median (IQR) | 67.4 (14.6) | 71.3 (11.9) | 3.41e-05 | 75.9 （13.9） | 1.30e-32 |
| sex<br>-Women (%)<br>-Men (%) | <br>1225<br>892 | <br>103<br>81 | 0.62 | <br>277<br>205 | 0.87 |
| BMI<br>mean (SD) | 27.54 (4.26) | 27.77 (4.17) | 0.47 | 27.11 (3.95) | 0.04 |
| skin color<br>-Olive color-light brown<br>- White<br>- Pale | <br>309<br>1572<br>236 | <br>19<br>147<br>18 | 0.45 | <br>48<br>386<br>48 | 0.17 |
| hair color when young<br>-Black<br>-Brown/dark blonde<br>-Light blonde<br>-Red | <br>124<br>154<br>396<br>57 | <br>8<br>140<br>28<br>8 | 0.56 | <br>25<br>340<br>105<br>12 | 0.12 |
| eye color<br>-Brown<br>-Intermediate<br>-Blue | <br>474<br>171<br>1472 | <br>37<br>18<br>129 | 0.66 | <br>94<br>34<br>354 | 0.10 |
| pigment status<br>-Light<br>-Intermediate | <br>431<br>1195 | <br>33<br>113 | 0.98 | <br>108<br>278 | 0.11 |

| | | | | | |
|---|---|---|---|---|---|
| -Dark | 491 | 38 | | 96 | |
| baldness (baldness_baseline_cat)<br>-No/almost no<br>-Mild<br>-Severe | 1570<br>291<br>256 | 120<br>28<br>36 | 2.32e-3 | 314<br>78<br>90 | 1.41e-05 |
| number of naevi<br>- >=100<br>- 50-99<br>- 25~49<br>- <25 | 41<br>107<br>297<br>1672 | 2<br>9<br>22<br>151 | 0.26 | 7<br>18<br>63<br>394 | 0.24 |
| Glogau wrinkle classification<br>- 1 and 2<br>- 3<br>- 4 | 149<br>1674<br>294 | 9<br>139<br>36 | 0.02 | 16<br>336<br>130 | 6.78e-13 |
| socioeconomic status<br>-high<br>-medium<br>-low | 616<br>1234<br>267 | 50<br>105<br>29 | 0.29 | 116<br>282<br>84 | 1.90e-03 |
| history of living in a sunny country<br>-Yes<br>-No | 122<br>1995 | 15<br>169 | 0.19 | 39<br>443 | 5.57e-02 |
| Tendency to develop sunburn<br>-Yes<br>-No | 693<br>1424 | 61<br>123 | 0.91 | 179<br>303 | 0.06 |
| alcohol intake<br>g/day<br>mean (SD) | 7.81 (8.05) | 7.96 (9.72) | 0.81 | 6.95 (6.78) | 0.03 |
| smoking<br>-Ever<br>-Never | 1429<br>688 | 123<br>61 | 0.86 | 314<br>168 | 0.32 |
| coffee consumption (cups/day) median (SD) | 3.03 (1.97) | 3.01 (1.97) | 0.92 | 2.57 (1.65) | 2.20e-06 |

| | | | | | |
|---|---|---|---|---|---|
| GRS_KC, median (SD) | 1.03 (0.25) | 1.04 (0.25) | 0.50 | 1.08 (0.29) | 2.38e-04 |
| GRS_MM, median (SD) | 7.03 (0.48) | 7.04 (0.45) | 0.83 | 7.09 (0.50) | 0.02 |

Right-censored samples: Never had skin cancer before the end of the study;
Left-censored: Diagnosed with skin cancer prior to the facial photo being taken;
Event: Diagnosed with skin cancer at least 30 days after the facial photo was taken.
FBSE: Full-body skin examination;
GRS_KC: Polygenetic risk score for KC;
GRS_MM: Polygenetic risk score for melanoma

#### 2. Survival analysis prediction c-index based on imputed or non-imputed risk factors.

Table S2: Comparison of c-index (mean/SD) between imputed and non-imputed risk factors.

| Method (predictors) | Imputed<br>N_event = 228<br>N_right-censored = 537 | Non-imputed<br>N_event = 184<br>N_right-censored= 434 |
|---|---|---|
| CPH (age only) | 0.550/0.048 | 0.559/0.043 |
| CPH (18 known risk factors) | 0.589/0.034 | 0.593/0.044 |
| DCPH (18 known risk factors) | 0.572/0.044 | 0.575/0.061 |
| CPH (facial endophenotypes) | 0.685/0.033 | 0.680/0.043 |
| DCPH (facial endophenotypes) | 0.721/0.045 | 0.719/0.045 |
| DCPH (facial endophenotypes + age) | 0.723/0.039 | 0.721/0.045 |
| DCCPH 2D facial image without extracted endophenotypes | 0.713/0.041 | 0.711/0.051 |

CPH: Cox proportional hazard regression;
DCPH: Deep cox proportional hazard regression;
DCCPH: Deep convolutional cox proportional hazards regression;
Only age-matched right-censored samples with > 6 follow-years were included in this analysis;
Events included skin cancer on any location, with imputed covariates (N=228) and non-imputed (N=184).

#### 3. Survival analysis based on all right-censored samples, or right-censored sample > 6 follow-up years.

| | |
|---|---|
| model | lifelines.CoxPHFitter |
| duration col | 'duation' |
| event col | 'event' |
| baseline estimation | breslow |
| number of observations | 2810 |
| number of events observed | 228 |
| partial log-likelihood | -1712.58 |
| time fit was run | 2023-04-07 15:04:12 UTC |

| | coef | exp(coef) | se(coef) | coef lower 95% | coef upper 95% | exp(coef) lower 95% | exp(coef) upper 95% | z | p | -log2(p) |
|---|---|---|---|---|---|---|---|---|---|---|
| age | 0.05 | 1.05 | 0.01 | 0.03 | 0.06 | 1.03 | 1.07 | 5.22 | <0.005 | 22.40 |
| sex | 0.07 | 1.07 | 0.17 | -0.26 | 0.39 | 0.77 | 1.48 | 0.39 | 0.69 | 0.53 |
| hair color | -0.13 | 0.88 | 0.16 | -0.45 | 0.19 | 0.63 | 1.21 | -0.80 | 0.43 | 1.23 |
| glogau | 0.13 | 1.14 | 0.16 | -0.18 | 0.45 | 0.84 | 1.57 | 0.84 | 0.40 | 1.32 |
| skin color | -0.01 | 0.99 | 0.15 | -0.30 | 0.27 | 0.74 | 1.31 | -0.10 | 0.92 | 0.12 |
| smoking | -0.01 | 0.99 | 0.15 | -0.30 | 0.29 | 0.74 | 1.33 | -0.05 | 0.96 | 0.06 |
| eye color | -0.06 | 0.94 | 0.16 | -0.38 | 0.26 | 0.68 | 1.29 | -0.38 | 0.70 | 0.51 |
| BMI | 0.00 | 1.00 | 0.02 | -0.03 | 0.03 | 0.97 | 1.03 | 0.18 | 0.86 | 0.22 |
| easily sunburn | -0.16 | 0.85 | 0.15 | -0.46 | 0.13 | 0.63 | 1.14 | -1.08 | 0.28 | 1.84 |
| naevi | -0.02 | 0.98 | 0.09 | -0.20 | 0.15 | 0.82 | 1.16 | -0.28 | 0.78 | 0.35 |
| alcohol intake | 0.00 | 1.00 | 0.01 | -0.01 | 0.02 | 0.99 | 1.02 | 0.44 | 0.66 | 0.60 |
| coffee consumption | 0.04 | 1.04 | 0.04 | -0.04 | 0.12 | 0.96 | 1.12 | 1.04 | 0.30 | 1.75 |
| pigment status | -0.22 | 0.81 | 0.31 | -0.83 | 0.39 | 0.44 | 1.48 | -0.70 | 0.49 | 1.04 |
| social economic status | 0.08 | 1.09 | 0.11 | -0.14 | 0.30 | 0.87 | 1.35 | 0.74 | 0.46 | 1.13 |
| baldness | -0.11 | 0.90 | 0.10 | -0.30 | 0.09 | 0.74 | 1.10 | -1.04 | 0.30 | 1.74 |
| GRS_KC | 0.28 | 1.33 | 0.26 | -0.23 | 0.80 | 0.79 | 2.22 | 1.08 | 0.28 | 1.85 |
| GRS_MM | 0.06 | 1.06 | 0.14 | -0.22 | 0.34 | 0.80 | 1.40 | 0.41 | 0.68 | 0.55 |
| sunny country history | -0.14 | 0.87 | 0.26 | -0.65 | 0.36 | 0.52 | 1.43 | -0.56 | 0.57 | 0.80 |

| | |
|---|---|
| Concordance | 0.64 |
| Partial AIC | 3461.17 |
| log-likelihood ratio test | 53.66 on 18 df |
| -log2(p) of ll-ratio test | 15.56 |

Figure S1: CPH model included all right-censored samples. Among 18 known risk factors, only age was found statistically significant. Age was not matched between events and right-censored samples.

| | |
|---|---|
| **model** | lifelines.CoxPHFitter |
| **duration col** | 'duation' |
| **event col** | 'event' |
| **baseline estimation** | breslow |
| **number of observations** | 895 |
| **number of events observed** | 228 |
| **partial log-likelihood** | -1484.60 |
| **time fit was run** | 2023-04-07 15:05:19 UTC |

| | coef | exp(coef) | se(coef) | coef lower 95% | coef upper 95% | exp(coef) lower 95% | exp(coef) upper 95% | z | p | -log2(p) |
|---|---|---|---|---|---|---|---|---|---|---|
| **age** | 0.03 | 1.03 | 0.01 | 0.01 | 0.05 | 1.01 | 1.05 | 2.70 | 0.01 | 7.19 |
| **sex** | 0.13 | 1.14 | 0.17 | -0.20 | 0.47 | 0.82 | 1.60 | 0.79 | 0.43 | 1.22 |
| **hair color** | -0.18 | 0.83 | 0.17 | -0.51 | 0.15 | 0.60 | 1.16 | -1.09 | 0.28 | 1.85 |
| **glogau** | 0.68 | 1.98 | 0.17 | 0.35 | 1.02 | 1.41 | 2.78 | 3.95 | <0.005 | 13.67 |
| **skin color** | -0.51 | 0.60 | 0.18 | -0.87 | -0.15 | 0.42 | 0.86 | -2.80 | 0.01 | 7.62 |
| **smoking** | 0.07 | 1.07 | 0.15 | -0.23 | 0.37 | 0.79 | 1.44 | 0.43 | 0.67 | 0.59 |
| **eye color** | -0.01 | 0.99 | 0.17 | -0.34 | 0.32 | 0.71 | 1.37 | -0.07 | 0.94 | 0.09 |
| **BMI** | 0.00 | 1.00 | 0.02 | -0.03 | 0.03 | 0.97 | 1.04 | 0.09 | 0.92 | 0.11 |
| **easily sunburn** | -0.16 | 0.85 | 0.15 | -0.46 | 0.14 | 0.63 | 1.15 | -1.03 | 0.30 | 1.72 |
| **naevi** | 0.06 | 1.06 | 0.09 | -0.11 | 0.24 | 0.89 | 1.27 | 0.69 | 0.49 | 1.02 |
| **alcohol intake** | -0.00 | 1.00 | 0.01 | -0.02 | 0.01 | 0.98 | 1.01 | -0.31 | 0.76 | 0.40 |
| **coffee consumption** | 0.06 | 1.06 | 0.04 | -0.02 | 0.13 | 0.98 | 1.14 | 1.45 | 0.15 | 2.78 |
| **pigment status** | -0.18 | 0.83 | 0.32 | -0.80 | 0.44 | 0.45 | 1.55 | -0.58 | 0.56 | 0.83 |
| **social economic status** | -0.14 | 0.87 | 0.11 | -0.36 | 0.08 | 0.70 | 1.08 | -1.23 | 0.22 | 2.20 |
| **baldness** | -0.05 | 0.95 | 0.10 | -0.25 | 0.16 | 0.78 | 1.17 | -0.45 | 0.65 | 0.61 |
| **GRS_KC** | 0.04 | 1.05 | 0.28 | -0.50 | 0.59 | 0.61 | 1.80 | 0.16 | 0.87 | 0.20 |
| **GRS_MM** | -0.10 | 0.91 | 0.15 | -0.39 | 0.20 | 0.67 | 1.23 | -0.63 | 0.53 | 0.91 |
| **sunny country history** | -0.15 | 0.86 | 0.26 | -0.67 | 0.36 | 0.51 | 1.43 | -0.59 | 0.56 | 0.84 |

| | |
|---|---|
| **Concordance** | 0.61 |
| **Partial AIC** | 3005.20 |
| **log-likelihood ratio test** | 50.03 on 18 df |
| **-log2(p) of ll-ratio test** | 13.71 |

Figure S2: CPH model only included right-censored samples > 6 follow-up years. Among 18 known risk factors, age, Glogau and skin color were found statistically significant. Age was not matched between events and right-censored samples.

### 4. Detailed architectures of the deep learning models used in this study

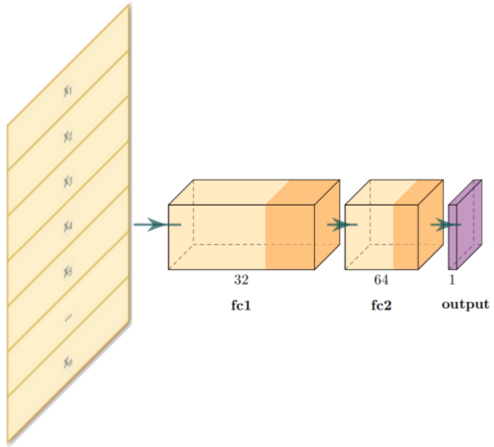


(a) Architecture of the DCPH model. $x_1 \sim x_n$ are n input variables (n=18 for risk factors or, n=200 for endophenotypes). There are 32 neurons in the first fully connected layer (fc1) and 64 neurons in the second fully connected layer (fc2). The DCPH model outputs a risk score in its output layer.

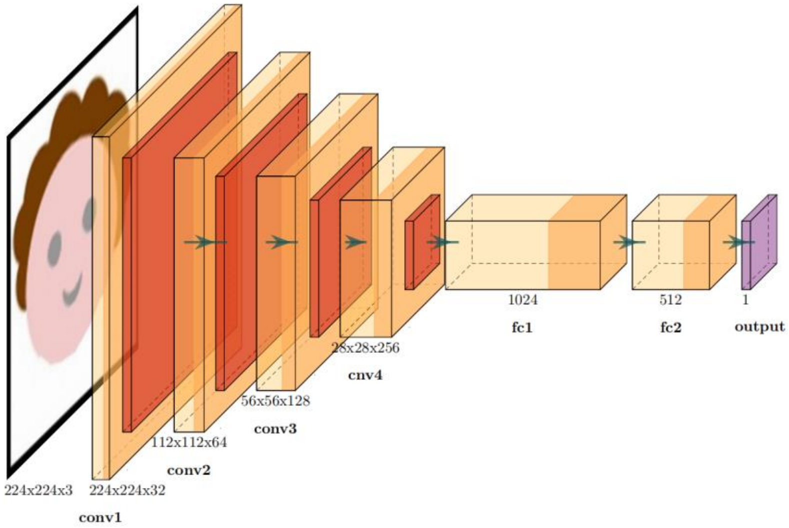


(a) Architecture of the DCCPH model. The size of the input 2D RGB facial image is 224x224x3, followed by 4 convolutional layers and 2 fully connected layers. The DCCPH model outputs a risk score in its output layer.

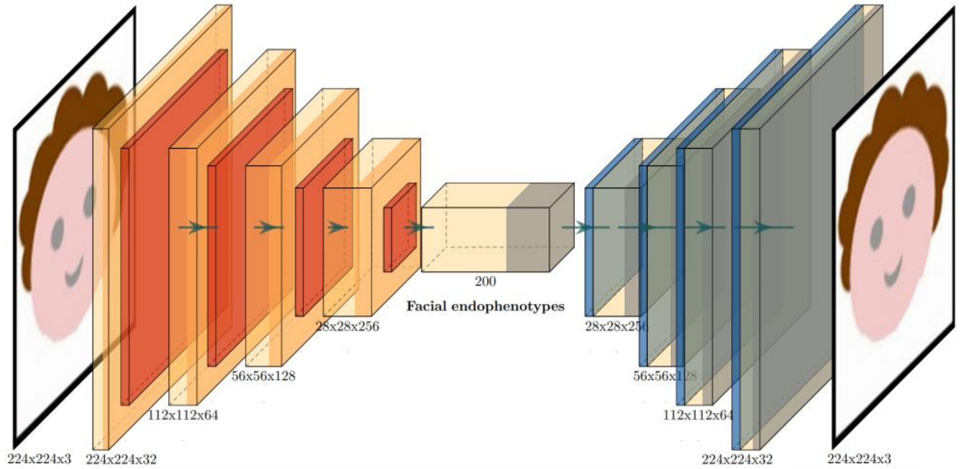


(c) Architecture of the autoencoder. There are 4 convolutional layers in the downsampling stage, while the upsampling process is a mirror process.

Figure S3: Detailed architectures of the a) DCPH model, b) DCCPH model, and c) autoencoder.

#### 5. Associations analysis between facial endophenotypes and skin cancer risk factors

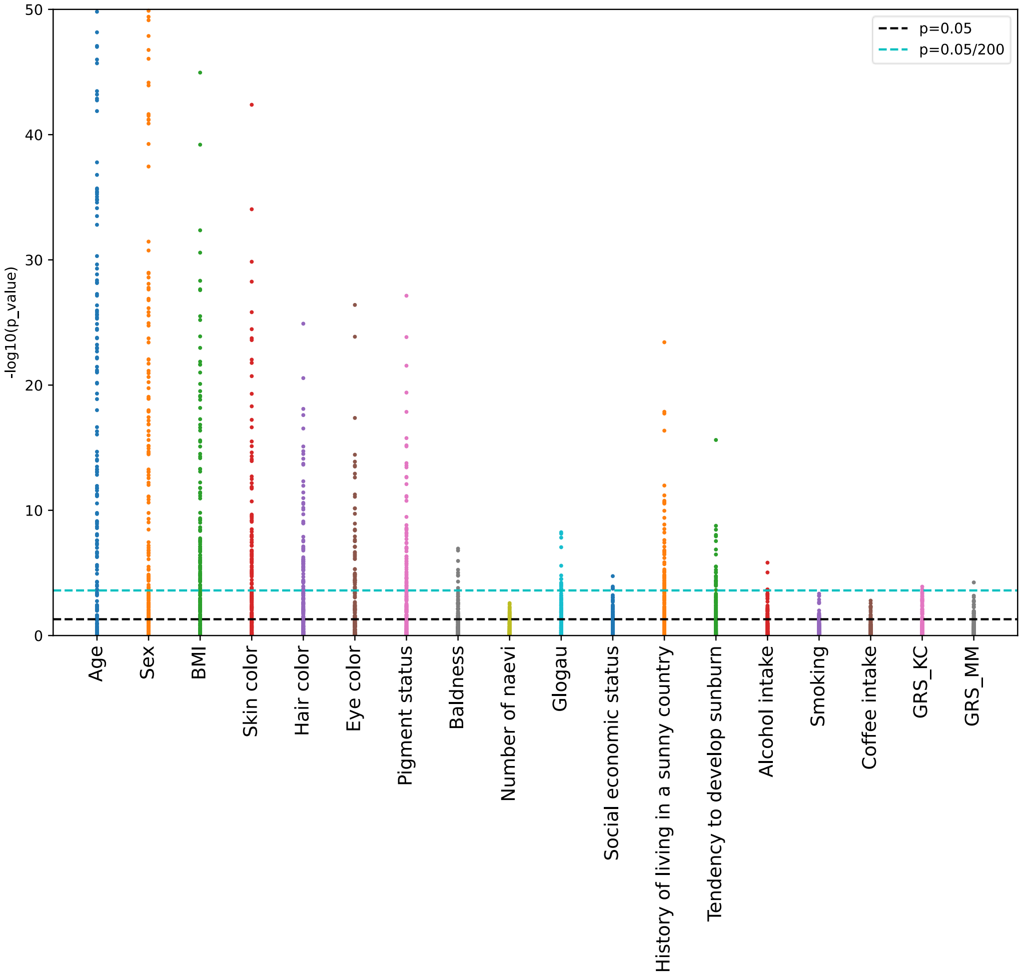


Figure S4: Associations between facial endophenotypes and risk factors, corrected for age and sex as confounders in linear regression analysis (N=3,371). Each point means an endophenotype, so there are 200 in each column.

#### 6. Implementation details of our explainable artificial intelligence (XAI) techniques

After the training of the autoencoder, each input facial image is represented by a datapoint (point d in Figure 5a) in the endophenotypes space. The endophenotype space (dimension: 200), derived from the autoencoder, can be viewed as a vector space. By selectively decoding a subset of endophenotypes, the decoder is able to perform sampling along a vector direction (which is determined by the weights of endophenotypes) and reconstruct a series of facial shapes showing facial deformation related to the combination of multiple endophenotypes (Figure S5a).

For example, when investigating the association between facial images and BMI, we firstly run linear regression for endophenotype $z_i$ as dependent variable:

$$z_i = \beta_i + \beta_i^{age} x_{age} + \beta_i^{sex} x_{sex} + \beta_i^{BMI} x_{BMI} \qquad \text{(S1)}$$

where i = 1, 2, …200 is the index of 200 endophenotypes, $\beta_i$ is the coefficient of the linear regression, $x_{BMI}$ is the independent variable, $x_{age}$ and $x_{sex}$ are covariates.

We select statistically significant endophenotypes, by setting $\beta_i^{BMI} = 0$ if the corresponding p-value (after correction for multiple testing) $p_i > 0.5$. A vector $\vec{v}$ is then determined by 200 coefficients:

$$\beta_i^{BMI} = 0, \text{ if } p_i > 0.5, i=1,2, ..., 200;$$

$$\vec{v} = \beta_1^{BMI} * z_1 + \beta_2^{BMI} * z_2 + ... \beta_{200}^{BMI} * z_{200}, i= 1, 2, …, 200 \qquad \text{(S2)}$$

By sampling and reconstructing facial images along such a vector direction, we are able to generate a series of facial images corresponding to the changes of BMI (Figure S5a).

The range of the sampling and decoding along the vector $\vec{v}$ was set as $\bar{d}$ ± 2*SD, where $\bar{d}$ is the mean of the data points in the study population, and SD is the standard deviation of L (Figure S5a). Point d' is the projection of d on vector $\vec{v}$, and L is the distance between d' and $\bar{d}$ (Figure S5a).

Similarly, when the vector direction is determined by coefficients from the survival analysis of skin cancer, we are able to visualize facial changes corresponding to the risk of developing skin cancer.

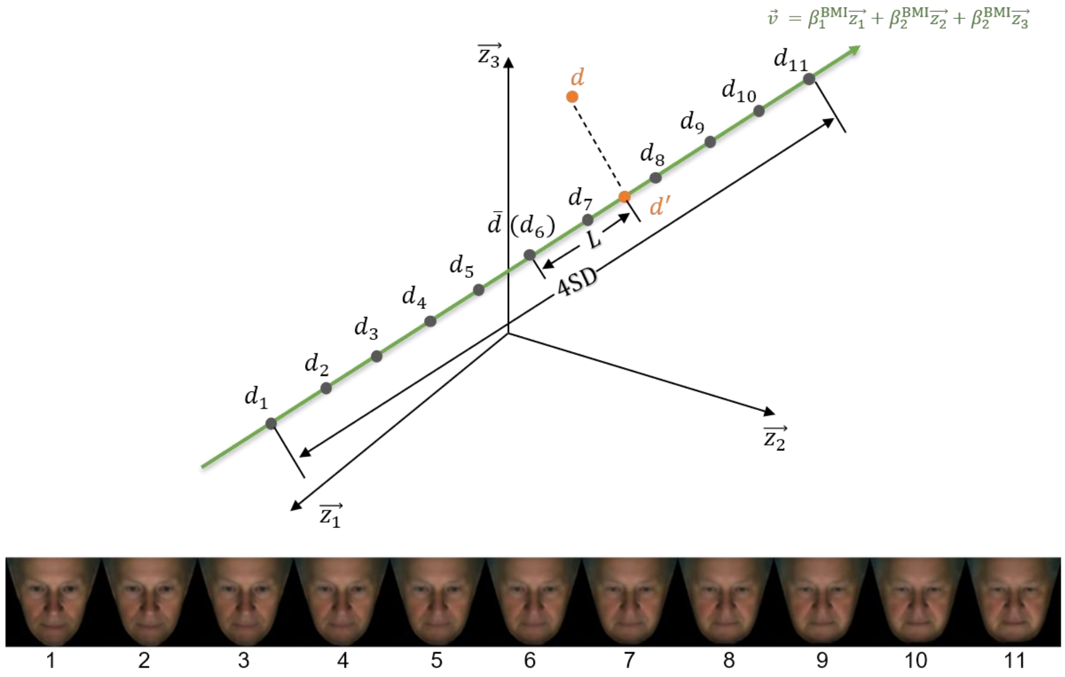


(a) An example of selectively decoding statistically significant endophenotypes in the linear regression analysis of BMI. Endophenotype space as a vector space with basis vectors $z_1$, $z_2$, ..., $z_{200}$ (only 3 dimensions are plotted in the figure). Vector $\vec{v}$ is determined by Equation (S1) and (S2). Point d is a datapoint of an input facial image, and d' is its projection onto vector $\vec{v}$. $\bar{d}$ is the mean of data points of all input facial images in the study population. L is the distance between $\bar{d}$ and d', and SD is the standard deviation of L among the study population. $d_1$, $d_2$, …, $d_{11}$ are sampled points along the vector $\vec{v}$, and facial images 1, 2, ..., 11 are reconstructed from $d_1$, $d_2$, ..., $d_{11}$ via the decoder.

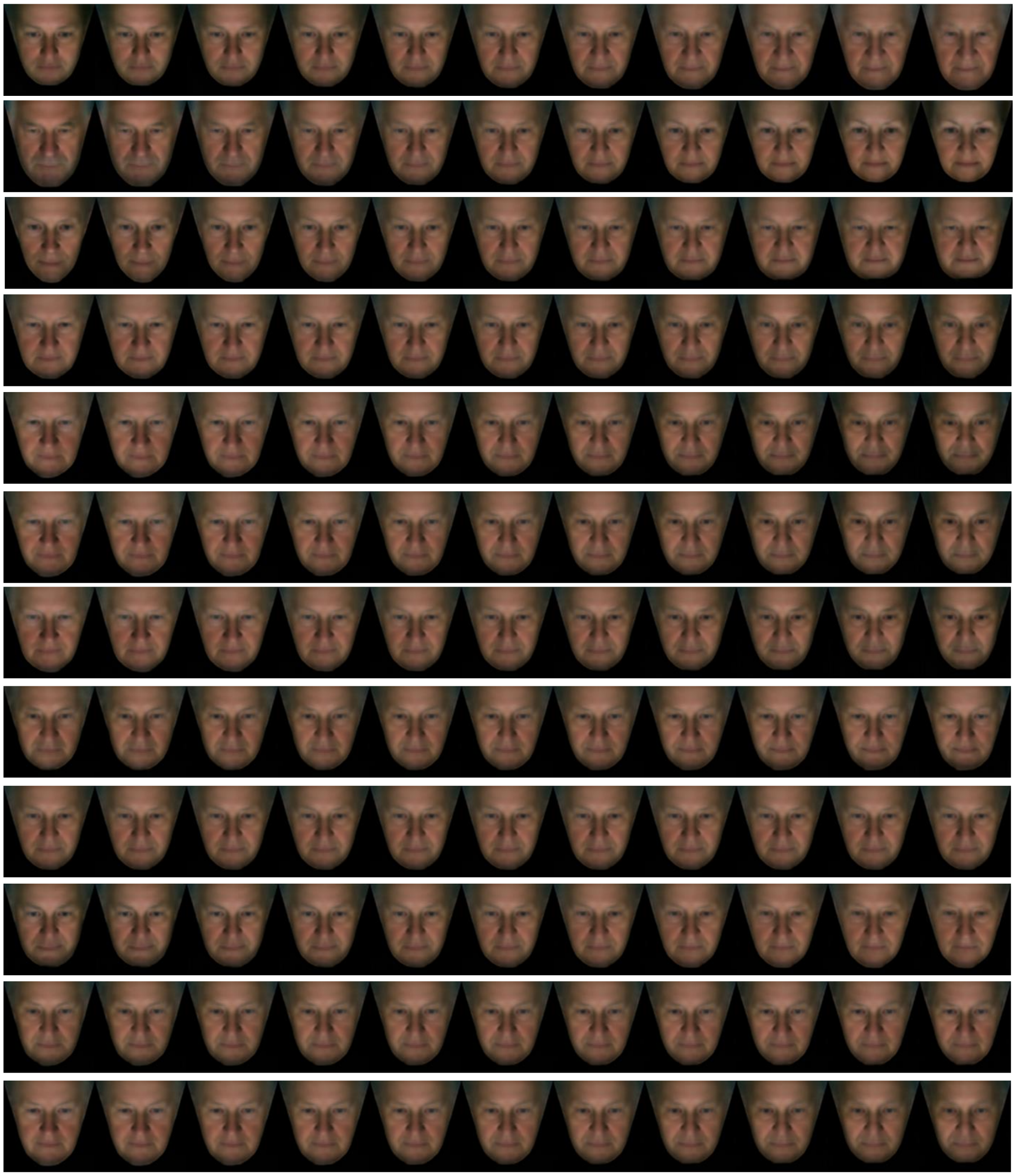
*Age
Young -> Old
*Sex
Male -> Female
*BMI
Low -> High
*Skin color
Light -> Dark
*Hair color
Light -> Dark
*Eye color
Blue -> Brown
*Pigment Status
Light -> Dark
*Baldness
No -> Severe
Number of Nevi
Low -> High
*Glogau
Low -> High
*Social Economic Status
Low -> High
*History of living in
a sunny country
No -> Yes

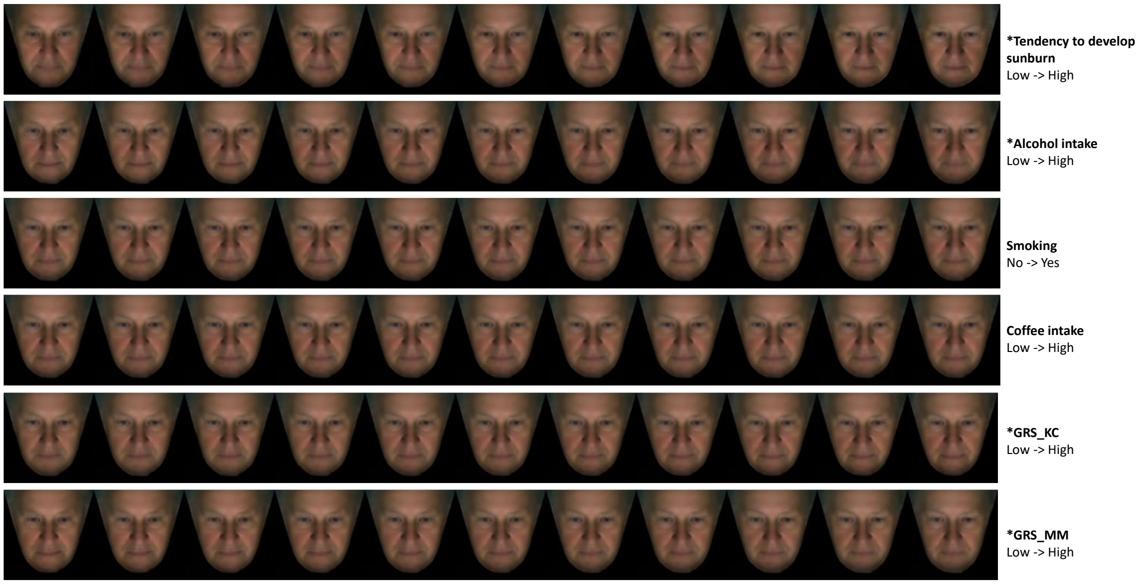


(b) Visualization results for the 18 risk factors in the association analysis (Figure S4). Risk factors with marker '*' means statistically significant.

Figure S5: Implementation details of the explainable AI (XAI) techniques. Facial images were reconstructed by decoding facial endophenotypes which were statistically significant in the analysis.

#### 7. Binary prediction between right-censored participants and participants ever diagnosed with skin cancer

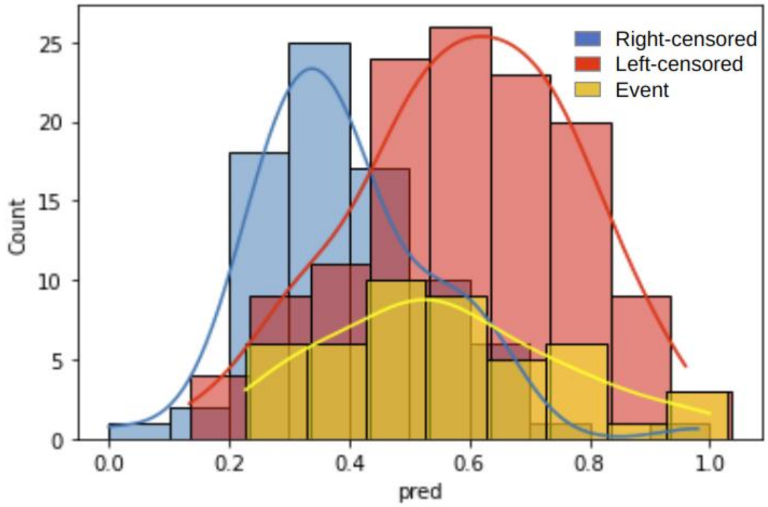


Figure S6: Binary prediction between right-censored participants (>6 follow-up years) and participants diagnosed with skin cancer (including both left-censored participants and events) based on facial endophenotypes. X-axis: prediction score normalized from 0 to 1; Y-axis: counts of the histogram. The AUC between right-censored data and left-censored data was 0.802, while the AUC between right-censored data and events was 0.747. Results were based on the test set.

#### 8. Distribution of the Time-to-event of events

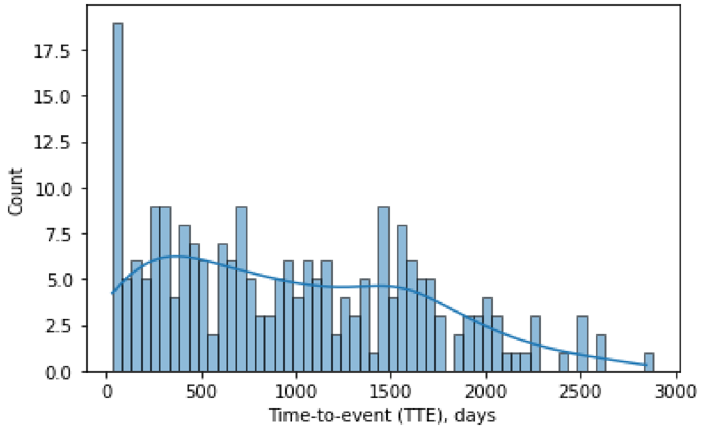


Figure S7: Histogram distribution of the Time-to-event, for events of skin cancer on any location (N=228).

#### 9. Preprocessing of the 2D facial images

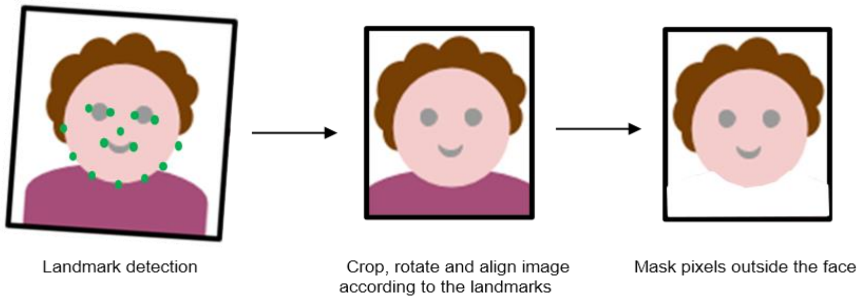


Figure S8: Preprocessing of the 2D facial images. Based on the locations of landmarks, the input image was firstly cropped, rotated and aligned into a standard position. Then, pixel regions outside the face were masked. In the end, only facial regions will be used for further analysis.

##### 10. Relationship between risk prediction score and time to event, for skin cancer at the body other than the face

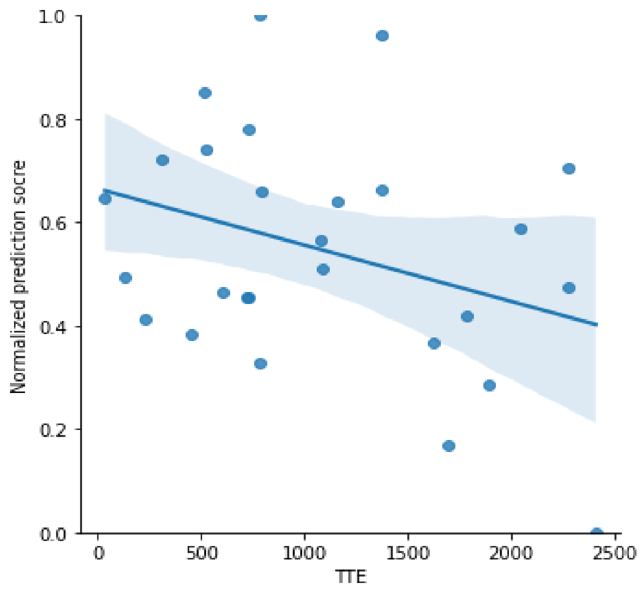


Figure S9: Relationship between risk prediction score and time to event (TTE) for N=27 events in the test set, in the analysis of (DCPH facial endophenotypes) for skin cancer at the body other than the face (Table 2). The X-axis represents the TTE in days, while the Y-axis represents the prediction risk score normalized to a range of 0 to 1. A higher predicted score indicates a higher risk of skin cancer.
